# A Novel Autosomal Dominant Childhood-Onset Disorder Associated with Pathogenic Variants in *VCP*

**DOI:** 10.1101/2023.06.19.23291232

**Authors:** Annelise Y. Mah-Som, Jil Daw, Diana Huynh, Mengcheng Wu, Benjamin C. Creekmore, William Burns, Steven A. Skinner, Øystein L. Holla, Marie F. Smeland, Marc Planes, Kevin Uguen, Sylvia Redon, Tatjana Bierhals, Tasja Scholz, Jonas Denecke, Martin A. Mensah, Henrike L. Sczakiel, Heidelis Tichy, Sarah Verheyen, Jasmin Blatterer, Elisabeth Schreiner, Jenny Thies, Christina Lam, Christine Spaeth, Loren Pena, Keri Ramsey, Vinodh Narayanan, Laurie H. Seaver, Diana Rodriguez, Alexandra Afenjar, Lydie Burglen, Edward B. Lee, Tsui-Fen Chou, Conrad C. Weihl, Marwan S. Shinawi

## Abstract

Valosin-containing protein (VCP) is an AAA+ ATPase that plays critical roles in multiple ubiquitin-dependent cellular processes. Dominant pathogenic variants in *VCP* are associated with adult-onset multisystem proteinopathy (MSP) that presents with myopathy, bone disease, dementia, and/or motor neuron disease. Through GeneMatcher, we identified 13 unrelated individuals who carry novel heterozygous *VCP* variants (12 *de novo*, 1 inherited) associated with a childhood-onset disorder characterized by developmental delay, intellectual disability, hypotonia, and macrocephaly. Trio exome sequencing or multigene panel identified nine missense variants, two in-frame deletions, one frameshift, and one splicing variant. We performed *in vitro* functional studies and *in silico* modelling to investigate the impact of these variants on protein function. In contrast to MSP variants, most missense variants had decreased ATPase activity, and one caused hyperactivation. Other variants were predicted to cause haploinsufficiency, suggesting a loss-of-function mechanism. This is the first description of *VCP*-related neurodevelopmental disease presenting in childhood.

## INTRODUCTION

Valosin-containing protein (VCP), also known as p97, Cdc48, and Ter94 in other organisms, is an ubiquitously expressed AAA+ protein (ATPase associated with other activities) that facilitates protein degradation through the ubiquitin-proteasome and autophagy pathways (1-2). By controlling degradation of key signaling molecules and misfolded proteins, it plays a critical role in multiple cellular functions, including autophagy and lysosomal degradation (3-5), maintenance of mitochondria (6-8), DNA replication and repair (9-11), and stress response (12-13). Cryo-electron microscopy (cryo-EM) shows it forms a homohexamer (14), with each identical monomer composed of three major domains (N, D1, and D2). The N-terminal domain regulates co-factor/adaptor protein binding, controlling the localization of the hexamer and its functionality. D1 and D2 are ATPase domains; ATP hydrolysis leads to a conformational change in VCP’s N domain, inducing substrate binding and release, and unfolding of the substrate through a central pore to facilitate degradation or remodeling (1, 15, 16).

Heterozygous pathogenic variants in *VCP* (MIM #601023) are associated with multisystem proteinopathy (MSP), or inclusion body myopathy with early onset Paget disease and frontotemporal dementia (IBMPFD1) (MIM #167320). This autosomal dominant disorder presents in adulthood with incomplete penetrance of several phenotypes: inclusion body myopathy (proximal and distal muscle weakness), Paget disease of bone (localized bone deformity and pain), frontotemporal dementia (executive function deficits), and motor neuron disease (17-20). Over 250 patients have been reported in the literature. Pathogenic variants that cause MSP are localized at the N-D1 interface in three-dimensional space (**Figure 1**) and are hypothesized to alter co-factor binding or unfolding kinetics, which causes disease via altering stability of key signaling factors or causing accumulation of proteins that form inclusion bodies (21, 22).

**Figure 1:**
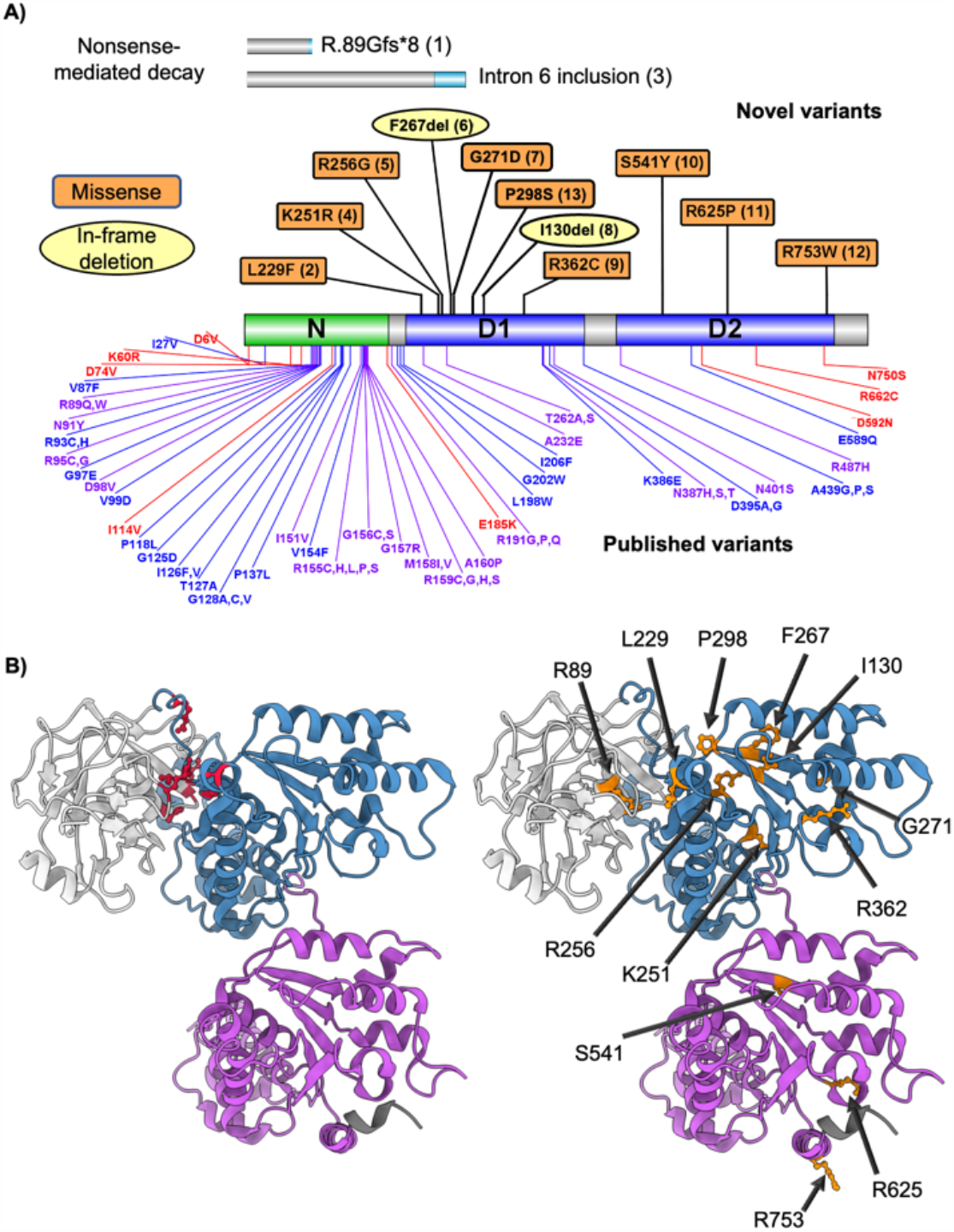
Novel variants in *VCP* include different sites and types of mutation than previously described in adult patients. A) Displayed is the canonical VCP protein and the N, D1, and D2 domains (59). The theoretical protein products for Probands 1 and 3 are shown above; novel sequence is in light blue, and both products are predicted to undergo nonsense-mediated mRNA decay before protein production. In-frame deletions are noted by yellow circles and missense mutations in our cohort are denoted in red squares, followed by the proband ID in parentheses. Below the bar, 44 previously described missense mutation sites are noted in blue (classic MSP with IBM, PDB, and/or FTD), red (any other phenotype), or purple (classic MSP + other phenotype overlap) (UniProt, 17, 40). B) Cryo-EM structures with a single subunit of VCP shown (60), with N domain in white, D1 in blue, and D2 in purple. On the left, marked in red are several classical MSP-associated variants, clustered in the N-D1 interface. On the right are the sites of variants described in this cohort, which are spread across the protein.

A growing number of pathogenic variants in *VCP*, some outside of the N-D1 interface, have been associated with other neurological conditions including isolated frontotemporal dementia (FTD)(23), familial amyotrophic lateral sclerosis (ALS)(24), Parkinson disease (25, 26), hereditary spastic paraplegia (27), Charcot-Marie-Tooth (CMT)(28), and autism (29). How these variants affect VCP function is not fully understood, but they may represent a neuron-predominant form of MSP.

Here, we describe a cohort of thirteen individuals with heterozygous *VCP* pathogenic variants including novel missense, in-frame deletion, frameshift, and splice variants, associated with a childhood-onset disorder characterized by developmental delay (DD), intellectual disability (ID), hypotonia, and macrocephaly.

## MATERIALS AND METHODS

### Standard Protocol Approvals, Registrations, and Proband Consents

GeneMatcher (61) facilitated the identification of probands with *VCP* variants. Written informed consent and authorization for publication was received from probands’ guardians using institution-specific protocols. Probands 1, 2, 3, and 6 signed a consent form approved by Washington University IRB (Media Authorization for the Use and Disclosure of Protected Health Information). Proband 9 signed a consent form approved by Seattle Children’s Hospital IRB (Media Authorization for the Use and Disclosure of Protected Health Information). Proband 11 signed a consent form approved by WCG IRB (Protocol #20120789) that included consent for publication of photographs. Proband 12 considered exempted from IRB requirements at their institution. Proband 13 signed a consent form approved by the Hospital Trousseau (Protocol #2010-A00715-34). No interventions were performed on patients and no biological specimens were collected from participants for this research study.

### Genetic testing

Next generation sequencing was performed at each institution on a clinical basis for evaluation of specific clinical findings in all probands. See **Supplementary Materials** for details of testing on each proband. Briefly, *VCP* variants were identified on trio exome sequencing (ES) for all probands except Proband 5, who had an intellectual disability gene panel run at Brest University Hospital, and Proband 10, whose variant was identified via GeneDx’s Autism/ID Xpanded panel (Gaithersburg, MD). ES was analyzed by the following laboratories: GeneDx (Probands 2, 3, 9, 11, 12), Greenwood Genetic Center (Proband 1), Telemark Hospital (Proband 4), Institute of Human Genetics Hamburg (Proband 6), Charité Universitätsmedizin Berlin (Proband 7), and D&R Institute of Human Genetics Graz (Proband 8). All *VCP* variants were *de novo* except in Proband 12, where the variant was paternally inherited. All were reported by the performing laboratories as variants of uncertain significance (VUS).

### Clinical history

Demographic data, clinical and developmental histories, and results of additional diagnostic work-up were obtained from evaluation of the probands by their geneticist and/or neurologist, chart review, and/or gathering information from the proband’s parents (available in **Supplementary Materials**). All probands were examined by a clinical geneticist and their genetic results were discussed by a geneticist or a genetic counselor.

### In silico analysis

Variants were analyzed *in silico* using CADD and REVEL scoring (62, 63) and classified based on current ACMG criteria (33-35). mRNA and genomic sequences were acquired from NCBI Gene, and the reference sequences were NG_007887.1 (GRCh38.p13 Primary Assembly), NM_007126.5 (transitional endoplasmic reticulum ATPase isoform 1, VCP-201), and NP_009057.1. NCBI BLAST was used to assess conservation at the affected residues in *Homo sapiens, Pan troglodytes, Mus musculus, Danio rerio, Drosophila melanogaster*, and *Saccharomyces cerevisiae*. Sequences were manually annotated and manipulated in A plasmid Editor (ApE) v3.0.8 by M. Wayne Davis (jorgensen.biology.utah.edu/wayned/ape/). UnitProt (uniprot.org) was used to visualize protein structures; **Supplementary Figure 1** features PDB structure 7BP9 (64). The RaptorX Contact Prediction Server was used to analyze folding of the in-frame deletion variants (raptorx.uchicago.edu) (37, 38). **Figure 1A** was made using Illustrator for Biological Sequences (65). **Figure 1B** was made using ChimeraX 1.4 using PDB structure 5FTM, although variants were also examined using 5FTN (down vs up configuration of N domain)(60).

### In vitro ATPase activity assays

Human VCP plasmid (TCB197) was subjected to site-directed mutagenesis with primers containing mutations to create each of the indicated variants. Proteins were purified as described from *E. coli* (66). Purified VCP (12.5 mL of 50 mM; final concentration in the reaction was 25 nM) was diluted in 20 mL of assay buffer [5 mL of 5x assay buffer A (1x = 50 mM Tris pH 7.4, 20 mM MgCl2, 1 mM EDTA), mixed with 15 mL water and 25 mL 0.5M TCEP, 25 mL 10% Triton] to make the enzyme solution. 40 mL of enzyme solution was dispensed into each well of a 96 well plate. The ATPase assay was carried out by adding 10 mL of 1000 mM ATP (Roche, pH 7.5) to each well and incubating the reaction at room temperature for 25 min. Reactions were stopped by adding 50 mL of BIOMOL Green reagent (Enzo Life Sciences). Absorbance at 635 nm was measured after 4 min on the Synergy Neo Microplate Reader (BioTek). All assays were performed in triplicate and the activity was averaged from independent experiments.

### Statistical analysis

Data were expressed as number and percentage for categorical variables and as mean ± standard deviation (SD) or standard deviation/Z-score for quantitative ones. Statistical analysis was performed using the program IBM SPSS statistics, version 25. For ATPase assays, statistical significance was defined using a 2-way ANOVA across all samples compared to the WT control.

### Cell culture and immunoblotting

U2OS cells were transfected with pcDNA3.1 with mouse VCP (100% homologous to human at amino acid level) containing an in-frame c-terminal myc/his tag. Variants were made using site-directed mutagenesis with primers containing mutations to create each of the indicated variants. Cell transfection was performed using Lipofectamine 2000 (ThermoFisher) according to the manufacturer’s instructions with 1 µg of plasmid. The transfection complex was removed and the cells were replaced with new media after 24 h. After 48 hours, U2OS cells were suspended in radioimmunoprecipitation assay lysis buffer (50 mM Tris-HCl, pH 7.4, 150 mM NaCl, 1% NP-40 [Sigma, I3021], 0.25% Na deoxycholate [Sigma-Aldrich, 30970], 1 mM EDTA) supplemented with protease inhibitor cocktail (Sigma-Aldrich, S8820). Lysates were centrifuged at 14,000g for 10 min. Aliquots of the supernatant were solubilized in Laemmli sample buffer and equal amounts were separated on 10% SDS-PAGE gels, transferred to nitrocellulose, and blocked with 5% nonfat dry milk in TBST (Tris-buffered saline, 0.1% Tween 20 [Sigma-Aldrich, 30970]). Membrane was incubated with primary antibody at 1:500 dilution overnight followed by incubation with secondary antibody conjugated with horseradish peroxidase at a 1:5,000 dilution. The Amersham ECL Western Blotting Detection Reagents Kit (GE Healthcare, RPN3244) was used for protein detection, and immunoblots were visualized with G:Box Chemi XT4, Genesys version 1.1.2.0 (Syngene, Cambridge, UK). Antibodies used: anti-GAPDH (Cell Signaling Technology, 2118), anti-VCP (Fitzgerald, 10R-P104A), anti-MYC (Cell Signaling Technology, 2276).

## Data availability

We, or the sequencing laboratory, have submitted these variants and associated phenotypes to ClinVar. Anonymized data will be shared by request from any qualified investigator.

## RESULTS

We describe thirteen probands with novel heterozygous *VCP* variants associated with childhood-onset neurodevelopmental disease. The probands ranged from 2 to 22 years old (mean 11 ± 6), came from multiple ethnic backgrounds, and shared phenotypic features that have not been previously associated with *VCP*-related disease including DD, ID, hypotonia, neurobehavioral abnormalities, dysmorphic features, and macrocephaly. Although their variants were classified as variants of uncertain significance (VUS) by the diagnostic laboratories, our *in vitro* and *in silico* analyses support that these variants are pathogenic and may represent a novel mechanism of disease for *VCP*.

### Novel variants in *VCP*

Thirteen *VCP* variants were identified on trio exome sequencing or gene panel: nine missense, two in-frame deletions, one frameshift leading to early termination, and one splice variant (**Table 1)**. All variants arose *de novo* except for Proband 12, who inherited the variant from his father. These variants contribute significant diversity to previously reported *VCP* genotypes. While most *VCP* variants linked to MSP are located at 37 residues at the N-D1 interface in the protein’s tertiary structure (20, 30, 31), the variants in our cohort spanned the entire length of the protein, including residues within D1 and D2 domains (**Figure 1**). One variant had been described in 2 relatives with adult-onset FTD (c.801_803del) (32), but all other variants had not been previously reported as pathogenic or benign variants.

**Table 1:** Thirteen variants in *VCP* associated with childhood-onset disease. Twelve of the variants are novel. The mRNA and protein coordinates are mapped to isoform 1 of *VCP* (NM_007126.5, NP_009057.1). For frameshift and splice variants, nonsense-mediated decay is predicted for transcripts due to premature stop codons before the last exon. D = domain of VCP protein. ACMG criteria include the following (33-35): inheritance mode (PS2 = pathogenic strong for *de novo* inheritance with maternity and paternity confirmed by exome, BS4 = benign strong for lack of segregation in family), population or allele frequency in gnomAD and other databases (PM2 = pathogenic moderate, absent from controls), and variant effect. Variant effects are scored differently based on type, for example for frameshift variants, PVS1-M = initially pathogenic very strong, corrected to moderate as haploinsufficiency is not known mechanism of disease. For missense variants, *in silico* prediction algorithms are used (PP3 = pathogenic supportive for multiple computational models predicting a deleterious effect, reweighted to moderate PP3-M or strong PP3-S based on degree of deleteriousness, PP2 = pathogenic supportive for missense variant in gene with low benign missense rate/missense variants are a known mechanism of disease). We give the final ACMG classification based on these criteria as LP = likely pathogenic, pathogenic, or VUS = variant of uncertain significance.

| ID | mRNA | Protein | D | ACMG Criteria |  |  |  |
| --- | --- | --- | --- | --- | --- | --- | --- |
|  |  |  |  | Inheritance | Pop freq | Effect | Class. |
| 1 | c.265del | R89Gfs*8 | N | De novo (PS2) | N/A (PM2) | PVS1-M | LP |
| 2 | c.685C>T | L229F | D1 | De novo (PS2) | N/A (PM2) | PP3 / PP2 | LP |
| 3 | c.709-2A>G | Splice variant | D1 | De novo (PS2) | N/A (PM2) | PVS1-M | LP |
| 4 | c.753G>T | K251N | D1 | De novo (PS2) | N/A (PM2) | PP3-M / PP2 | LP |
| 5 | c.766C>G | R256G | D1 | De novo (PS2) | N/A (PM2) | PP3-M / PP2 | LP |
| 6 | c.801_803del | F267del | D1 | De novo (PS2) | N/A (PM2) | PM4 | LP |
| 7 | c.812G>A | G271D | D1 | De novo (PS2) | N/A (PM2) | PP3-S / PP2 | LP |
| 8 | c.901_903del | I301del | D1 | De novo (PS2) | N/A (PM2) | PM4 | LP |
| 9 | c.1084C>T | R362C | D1 | De novo (PS2) | N/A (PM2) | PP3-S / PP2 | Path. |
| 10 | c.1622C>A | S541Y | D2 | De novo (PS2) | N/A (PM2) | PP3-M / PP2 | LP |
| 11 | c.1874G>C | R625P | D2 | De novo (PS2) | N/A (PM2) | PP3-S / PP2 | Path. |
| 12 | c.2257C>T | R753W | D2 | Paternal (BS4) | N/A (PM2) | PP3-M / PP2 | VUS |
| 13 | c.892C>T | P298S | D1 | De novo (PS2) | N/A (PM2) | PP3 / PP2 | LP |

Most probands underwent extensive genetic and metabolic testing prior to discovery of these variants (**Supplementary Table 2**), but no proband met diagnostic criteria for an alternative genetic condition. More importantly, our patients did not have clinical or laboratory features suggestive of MSP. The similar phenotypes and recurring clinical features as well as lack of other genetic abnormalities amongst the probands in this cohort strongly support a pathogenic role for heterozygous *VCP* variants in childhood-onset neurodevelopmental disease.

### In silico characterization of VCP variants

All variants were classified by their reporting laboratories as VUS. However, by applying ACMG criteria (33-35) and inheritance mode, we were able to reclassify all variants as “likely pathogenic” or “pathogenic” except for Proband 12, as it is unclear whether his father who shares the variant is affected (**Table 1, Supplementary Table 1**).

Two variants were predicted to lead to nonsense-mediated mRNA decay: a frameshift causing early termination in exon 3 in Proband 1 (R89Gfs*8) and a splice variant in Proband 3 (c.709-2A>G) leading to inclusion of intron 6 and early termination (**Figure 1A**). No transcript isoforms containing this intron were found by RNA-seq in the HAVANA project/Ensembl database. Interestingly, Proband 7’s c.812G>A variant is at the 3’ splice acceptor site of intron 7-exon 8, but splicing predictions were mixed (unaffected by SpliceAI, deleterious by dbscSNV)(36, 37). The presence of variants that are predicted to cause nonsense-mediated decay highly suggests that haploinsufficiency of VCP is a pathomechanism for this novel disease entity.

It is unclear what effect the in-frame deletions in Probands 6 and 8 would have on protein function. Proband 6’s F267del variant was recently reported in two relatives with FTD/aphasia diagnosed around age sixty, which suggests pathogenicity for adult-onset disease (32). Proband 8’s I301del variant had not been previously reported as pathogenic or benign in any genome databases. Based on cryo-EM structure, F267del and I301del remove residues from the center of a beta strand in the same pleated sheet in D1 (**Figure 1B**), relatively near the ATP binding site. Contact prediction using the deep-learning software RaptorX (38, 39) predicts that the beta sheet secondary structure would be retained in both cases without striking changes in the orientation of nearby alpha helices (**Supplementary Figure 1**). However, minor conformational changes could still interfere with ATP binding, hexamerization, protein dynamics, or other functions.

*In silico* algorithms predicted deleterious effects for all nine novel missense variants in this cohort (**Table 1, Supplementary Table 1**). Variants at these sites had not been reported in human disease. The residues involved are highly conserved between species, from human to *S. cerevisiae* (**Supplementary Figure 2**). Proband 4’s K251N variant affects the “Walker A motif” required for ATP interaction, so it likely has decreased ATPase activity (40, 41). AAA+ ATPases also have an arginine finger required to stabilize the leaving group; this is hypothesized to be R359 (42), although R256 (replaced by glycine in Proband 5) and R362 (replaced by cysteine in Proband 9) are also near the ATP binding pocket. Other notable mutations include L229F, in an alpha helix near the N-D1 interface, and two arginines (R625P and R753W), which sit at the proteasome-facing surface of the D2 ring (**Figure 1B**). Interestingly, four out of the nine missense variants affect arginines, a bulky and highly charged residue often involved in ionic interactions. ***In vitro* ATPase activity of variants**

**Figure 2:**
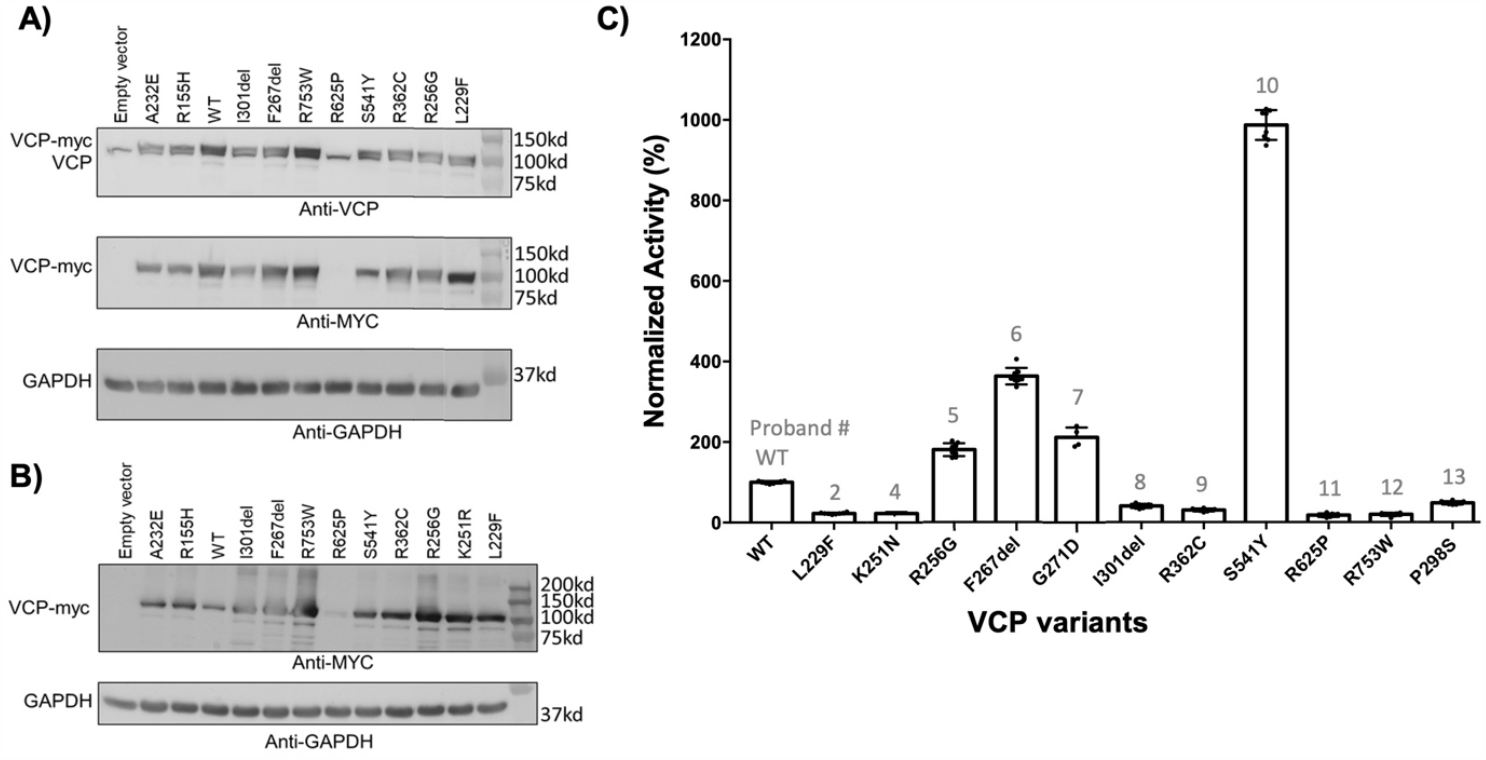
ATPase activity of novel *VCP* variants. A) Immunoblots of lysates from U2OS cells expressing empty vector or expression plasmids with *VCP* variants and a C-terminal Myc tag using an anti-VCP antibody (top), anti-Myc antibody (middle) or anti-GAPDH as loading control (bottom). VCP-WT and other variants include: classic MSP-associated variants A232E and R155H, L229F (Proband 2), R256G (Proband 5), F267del (Proband 6), I301del (Proband 8), R362C (Proband 9), S541Y (Proband 10), R625P (Proband 11), and R753W (Proband 12). Expression data for K251N (Proband 4) and G271D (Proband 7) were well-expressed and can be seen in Supplementary Figure 3. Note a small doublet is present with anti-VCP antibody, demonstrating the endogenous VCP (lower band) and the overexpressed myc-tagged VCP (upper band). The R625P variant fails to be expressed. B) Longer exposure of a similar set of lysates using the anti-myc antibody shows a faint band for the R625P variant, as well as high molecular weight smear and multiple degradation products for the novel variants. C) Recombinant purified human VCP protein was obtained from bacteria and *in vitro* ATPase activity was assessed using a standard colorimetric assay and normalized to wildtype VCP. Comparison of these variants against classic MSP mutations can be found in Supplementary Figure 3.

The frameshift and splice *VCP* variants predicted to lead to haploinsufficiency suggest that the other variants may similarly behave as loss-of-function alleles given our cohort’s shared phenotype. We first examined protein stability of each variant, expressed in U2OS cells. Most variants were expressed at similar levels to wildtype VCP, except for R625P (Proband 11) which had significantly decreased protein expression by Western blot (**Figure 2A**). There appeared to be accumulation of higher molecular weight species and smaller degradation products in this cohort’s variants, but not in the wildtype protein or MSP-associated variants (A232E, R155H) (**Figure 2B**). These data suggest that the missense variants reported here may destabilize VCP. We reasoned that these variants could cause loss of function by disrupting VCP’s intrinsic ATPase activity. We measured *in vitro* ATPase activity using recombinant human VCP protein purified from bacteria. As shown in previous publications, MSP-associated *VCP* variants increased ATPase activity, supporting a gain-of-function (**Supplementary Figure 3**, 22). Three of our variants fit this pattern: F267del, which was previously reported in adult-onset FTD (32) and had similar ATPase activity compared to classic MSP variants, and R256G and G271D, novel variants with intermediately elevated ATPase activity. In contrast, the other novel variants significantly decreased ATPase activity, with L229F, K251N, P298S, I301del, R362C, R625P and R753W having <50% of wildtype activity (**Figure 2C**). There was one hyperactivating variant, S541Y, which had ATP hydrolysis >1000% of wildtype and around twice that of MSP variants. This degree of ATPase activity may significantly affect VCP’s ability to coordinate ATP hydrolysis with co-factor association and protein unfolding.

**Figure 3:**
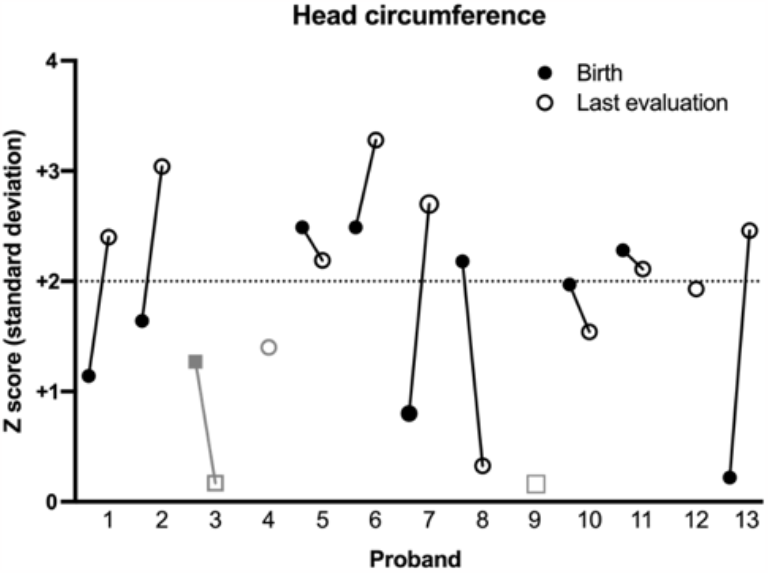
Facial features and head circumference. Macrocephaly is defined as head circumference >2 standard deviations (SD) above the mean. The number of SD (or Z score) was plotted for head circumference of each proband at birth (filled) and at last evaluation (open). Proband 4 (gray circle) has relative macrocephaly, as defined by standardized head circumference >2 SD above standardized height (43). Probands 3 and 9 (gray squares) were never macrocephalic. Proband 10 was evaluated at 98%ile, Z+1.97. Proband 12 was macrocephalic (>2 SD) at one point during his life, but not on last evaluation (SD +1.93).

### Clinical phenotypes

In our cohort of thirteen individuals (seven males and six females), ranging from two to twenty-two years of age at the time of last evaluation, the most striking phenotypic features were DD, ID, hypotonia, and macrocephaly (**Table 2, Supplementary Material**). There was also a significant number of probands with musculoskeletal abnormalities, abnormal but nonspecific brain MRI findings, psychiatric/behavioral disorders, ophthalmological abnormalities, and dysmorphic features.

**Table 2:**
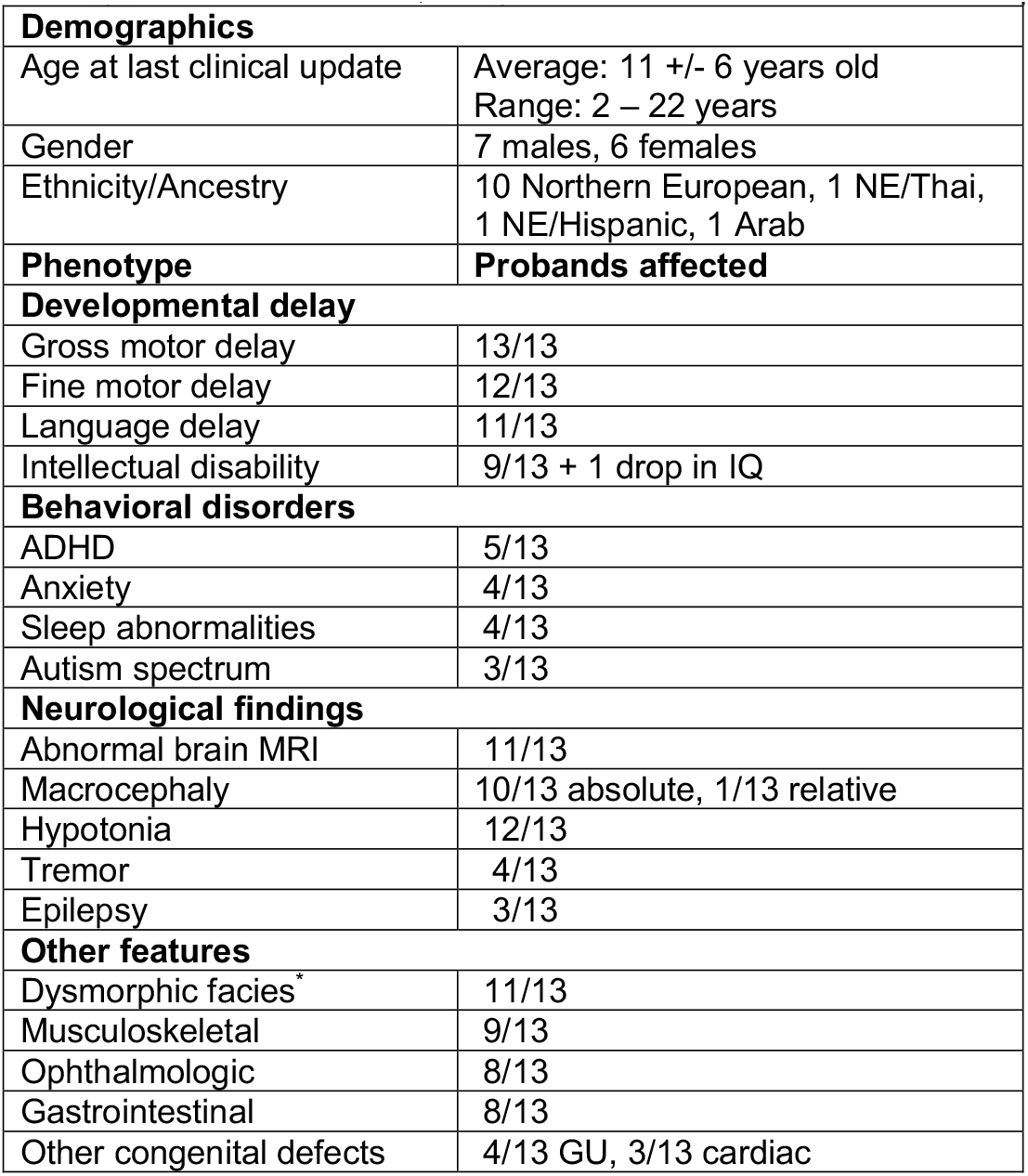
Summary of proband phenotypes. Phenotypes shared across probands with heterozygous *VCP* variants causing childhood-onset disease. Musculoskeletal abnormalities include kyphosis, scoliosis, hammer toes, pes cavus (including acquired), long bone deformities, torticollis, and hip dysplasia or misalignments; 5 probands required surgery, splinting, or orthotics. Ophthalmologic abnormalities include corneal clouding, astigmatism, strabismus, and hyperopia/myopia. Gastrointestinal features include pyloric stenosis, gastroesophageal reflux, constipation, and poor feeding. Genitourinary (GU) abnormalities include inguinal hernia, cryptorchidism, hydrocele, hypospadias, and duplicated collecting system. Cardiac abnormalities include ventricular septal defect, atrial septal defect/patent foramen ovale, and patent ductus arteriosus. ^*^ Includes patients with minor features.

| **Demographics** |  |
| --- | --- |
| Age at last clinical update | Average: 11 +/- 6 years old<br>Range: 2 – 22 years |
| Gender | 7 males, 6 females |
| Ethnicity/Ancestry | 10 Northern European, 1 NE/Thai, 1 NE/Hispanic, 1 Arab |
| **Phenotype** | **Probands affected** |
| **Developmental delay** |  |
| Gross motor delay | 13/13 |
| Fine motor delay | 12/13 |
| Language delay | 11/13 |
| Intellectual disability | 9/13 + 1 drop in IQ |
| **Behavioral disorders** |  |
| ADHD | 5/13 |
| Anxiety | 4/13 |
| Sleep abnormalities | 4/13 |
| Autism spectrum | 3/13 |
| **Neurological findings** |  |
| Abnormal brain MRI | 11/13 |
| Macrocephaly | 10/13 absolute, 1/13 relative |
| Hypotonia | 12/13 |
| Tremor | 4/13 |
| Epilepsy | 3/13 |
| **Other features** |  |
| Dysmorphic facies* | 11/13 |
| Musculoskeletal | 9/13 |
| Ophthalmologic | 8/13 |
| Gastrointestinal | 8/13 |
| Other congenital defects | 4/13 GU, 3/13 cardiac |

### Developmental delay & intellectual disability

The most striking shared phenotypes in this cohort were DD and ID, which had not been previously described in individuals with *VCP* variants. Specifics for each proband can be found in **Supplementary Table 3**. None of the probands had significant developmental regression.

All individuals in our cohort had either gross or fine motor delays, and most experienced both. Most gross motor delays were mild to moderate, with the majority learning to walk between eighteen months and three years old. Two probands had earlier delays but walked on time. Gross motor delays were associated with hypotonia, which was present in all but Proband 12. For example, Probands 4 and 8 required orthoses to walk, and Proband 4 uses a wheelchair. Hypotonia may have also affected fine motor development in our probands. For instance, Proband 2 currently uses a keyboard as generalized hypotonia makes handwriting difficult for him. Probands 11, 12, and 13 had tremor involving their hands which limited their fine motor skills. Additional findings that affected motor development include poor coordination or balance in Probands 5, 9, and 10, and hemiparesis with hemiatrophy in Proband 11.

Most individuals, except for Probands 1 and 12, exhibited language delay which was variable in severity. Five individuals were mostly nonverbal or spoke few words. Two had delayed speech onset, with first word around two years of age, but can now produce a reasonable vocabulary. Two others had onset of speech within normal range, but formation of sentences was delayed to four or five years old. Proband 13 had delayed onset of both first word and sentence formation, and her vocabulary remains limited. Other associated speech findings include dysarthria, apraxia, and expressive aphasia in one proband each.

Nine of thirteen individuals in this cohort exhibited ID, ranging from mild to severe. Few probands underwent formal IQ testing; three had IQs around 50, and two others were diagnosed with severe ID. Proband 12’s IQ was above the range for ID (<70), but he had regression of his IQ, testing at 105 initially and dropping to 76 six years later. This may have been related to the development of seizures, although cognitive decline due to the natural history of the condition cannot be excluded.

### Behavioral & psychiatric phenotypes

Six of thirteen individuals in this cohort were diagnosed with behavioral or psychiatric conditions, most with multiple diagnoses. This includes five diagnoses of ADHD, three formal diagnoses of autism, and four diagnoses of anxiety. Note that an individual with a frameshift variant in *VCP* was previously described to have autism; no further information was given but he may share features with this cohort (29). Several individuals had abnormal behaviors without a formal diagnosis, including head-banging behavior, hand-flapping, stereotypic hand movements, and aggressive outbursts (**Supplementary Table 4**). Four individuals had sleep difficulties.

### Neurological and neuroimaging findings

Twelve out of thirteen probands evaluated were noted to have hypotonia, making this a central phenotype in the cohort. Eleven had some abnormalities on brain MRI noted by the reporting radiologist (**Supplementary Table 4**). However, the findings were generally mild and nonspecific, and no significant brain malformations or structural anomalies were noted. Shared features included mild cerebral atrophy/generalized volume loss with prominence of CSF spaces (4/13), hydrocephalus with more significant cerebral atrophy (2/13), decreased white matter (3/13), and a thin corpus callosum (3/13). Proband 13 had the most abnormalities, with focal dysplasia in the left parietal cortex. Note that for most individuals, cerebral atrophy was generalized; however, Proband 11 had some mild atrophy of the frontal lobes that developed between childhood and adulthood, reminiscent of adult-onset MSP-related FTD. Probands 1 and 12, two of our least developmentally-affected probands, had normal MRIs.

Proband 12 had multiple seizures and was on antiepileptic medications for years. Two other individuals also had seizures, and Proband 4 had an epileptiform EEG but no clinical evidence of seizures. These three individuals (Probands 4, 5, and 10) were noted to have brain MRI abnormalities, but other probands with similar MRI changes did not have known seizures.

Ten individuals in our cohort had absolute macrocephaly, as defined by ≥98^th^ percentile head circumference at any point in their development, and one (Proband 4) had relative macrocephaly, as defined by standardized head circumference >2 SD above standardized height (43). The mechanisms of development of macrocephaly in our cohort appear to be varied. Six probands were macrocephalic at birth, while three others had head circumferences in the normal range at birth with progressive macrocephaly as they aged (**Figure 3**). In Probands 5 and 7, hydrocephalus may have contributed to enlargement of OFC pre- or postnatally. Proband 12’s mother also had a head circumference at the 98^th^ percentile, which may suggest familial factors for his macrocephaly.

### Evaluation for features of MSP

Few probands had testing to rule out MSP, as this is not a childhood-onset condition. Generally, there were no concerns for myopathy or Paget’s disease of the bone. All eight probands who had creatinine kinase (CK) measured were in the normal range. Nine probands had alkaline phosphatase (ALP) measured which were generally normal, although three had mild elevations during acute illness (**Supplementary Table 2**). The only proband to have a muscle biopsy was Proband 3, in whom no cytoplasmic inclusions were found. Proband 2 had a muscle ultrasound that was normal. As noted above, only Proband 11 showed MRI findings that could have been consistent with FTD, but clinical suspicion for the condition was low.

MSP is also associated with motor neuron disease, which can present with limb weakness, fasciculations, spasticity, and/or hyperreflexia (19). Most of our probands had low muscle tone, and Proband 12 had hyperreflexia. Testing for neuropathy or myopathy was performed in four probands based on their symptoms, and three had abnormal findings on electromyography (EMG) or nerve conduction velocity (NCV) testing. Proband 4 had a predominantly demyelinating sensorimotor polyneuropathy in the upper and lower extremities on NCV studies; his EMG was normal. Proband 9 initially had normal EMG and NCV; however repeat EMG four years later showed a mild sensorimotor polyneuropathy, and review of prior EMG showed similar findings in both. Proband 11 had an EMG that showed a mixed myopathic/neuronopathic picture, but repeat study six years later was normal (**Supplementary Table 2**). Overall, these results were not consistent with motor neuron disease seen in adult patients with MSP, but could be consistent with CMT (19). Only one additional proband had EMG testing (Proband 2), which was normal.

### Facial features

Eleven of thirteen individuals in our cohort had some dysmorphic facial features, although there was no obvious characteristic gestalt for this cohort (please contact corresponding author to request photographs if interested). The most common facial features included frontal bossing/prominent or broad forehead (9/13), thin upper lip (7/13), down-slanting palpebral fissures (5/13), low-set, posteriorly rotated, or dysplastic ears (4/13), nasal anomalies (4/13), deep set eyes (3/13), high/arched palate (3/13), and smooth philtrum (3/13) (**Supplementary Table 5**)

### Congenital anomalies

There were no common birth defects in our cohort, but most had at least one congenital anomaly (**Supplementary Table 5**). These included urogenital abnormalities such as cryptorchidism, hypospadias, and duplicated collecting system (5/13), cardiac malformations such as ventricular septal defect, patent ductus arteriosus, and patent foramen ovale (3/13), and musculoskeletal findings such as hip dysplasia and underdeveloped epiphyses (3/13). No cardiac findings required repair. Other findings included congenital torticollis, inguinal hernia, and umbilical hernia (each in one proband). Minor defects included palmar transverse crease, high foot arch, and hammer toes.

### Miscellaneous Findings

Eight out of the thirteen individuals in our cohort had ophthalmological abnormalities in infancy/childhood. Two developed strabismus, and one was noted to have punctate corneal clouding. Five probands had refractive errors including myopia, hyperopia and/or astigmatism. Eight probands had gastrointestinal abnormalities including five individuals with constipation and four with gastroesophageal reflux (**Supplementary Table 5**).

## DISCUSSION

We describe thirteen individuals with heterozygous variants in *VCP* (twelve *de novo*, one inherited) that cause a novel childhood-onset syndrome, distinct from adult-onset presentations, and characterized by DD, ID, hypotonia, and macrocephaly, with some behavioral/psychiatric disease and dysmorphic features. Functional studies and *in silico* analyses supported classification of twelve variants as pathogenic/likely pathogenic. Evidence supporting *VCP* pathogenic variants as the cause of this phenotype includes shared neurological and developmental features, extensive workup including genetic studies and brain imaging excluding other etiologies, and no alternative compelling diagnoses. The alterations in ATPase activity as demonstrated by *in vitro* studies suggested a different pathomechanism for this novel *VCP*-related disease compared to MSP, which could lead to earlier presentation of disease.

There is no clear genotype-phenotype correlation in our cohort. No link could be established between the severity of presentation, imaging findings, dysmorphic features, and the molecular consequence of the variant. For example, Probands 6 and 8 have in-frame deletions affecting residues in the same beta-pleated sheet, but have very different ATPase activity and exhibit differences in their MRI findings, facial features, and severity of their ID. Probands 5, 6, and 7, despite their variants having similar increases in ATPase activity, are discordant in head size, brain imaging, seizure history, and dysmorphic features. This implies that there is not a straightforward correlation between ATPase function and pathological consequence. Additionally, Probands 1 and 3, whose variants are predicted to lead to nonsense-mediated decay, do not have a more or less severe phenotype compared to probands with missense variants. However, there are clear differences in both the genotype and phenotype in our cohort compared to variants that cause MSP and other adult-onset *VCP*-related conditions.

*In-silico* predictions and *in vitro* data suggest a *VCP* loss-of-function mechanism via haploinsufficiency or decreased protein activity for these novel variants. Haploinsufficiency is suggested by Probands 1 and 3, whose variants are predicted to undergo nonsense-mediated mRNA decay and Proband 11’s R625P variant, which was poorly expressed in cells. *VCP* is predicted to be intolerant to loss of function variants (pLI ∼1, LOUEF 0.03) (gnomAD, 44). However, *in vivo* evidence supporting disease via haploinsufficiency of *VCP* is limited, as knock-out mice heterozygous for *VCP* were described as indistinguishable from their wildtype littermates (45). Studies focusing on neuron-specific knockout of *VCP* show more promising data, as knockdown of *VCP* by ∼60% decreased dendritic spine formation in cultured rodent neurons (46, 47), and neuron-specific knockout mice had reduced brain volume, hyperactivity, and poor spatial learning, which could correlate to our probands’ MRI and behavioral phenotypes (48). Further studies are needed to assess cognition and behaviors in heterozygous mice. In humans, 9p13 microdeletion syndrome leads to haploinsufficiency of VCP and is associated with DD, ID, and tremor, along with other findings such as short stature, genital anomalies, and precocious puberty (49-51). Although many other genes are present in the ∼2-5 Mb deletions, *VCP* should be considered a candidate gene for the 9p13 deletion neurological/developmental phenotype.

Most missense/in-frame deletion variants in our cohort displayed *VCP* loss-of-function via decreased ATPase activity. This is opposed to MSP-causing variants, in which ATPase function is intact or elevated (Supplementary Figure 3) (52-54). However, four variants (R256G, F267del, G271D, and S541Y) in our cohort increased ATPase activity. Proband 10’s S541Y hyperactivating variant with its super-rapid ATPase activity may not allow enough time for coordinated protein unfolding or co-factor binding and release. Other hyperactivating mutations, for example in *UBE3*A and *KIF1A*, have been described to cause disease when loss-of-function in these genes is the usual mechanism (55, 56). R256G and F267del in Probands 5 and 6 are near the N-D1 interface of VCP, and thus may alter adaptor or substrate binding similar to MSP-causing variants.

In fact, the c.801_803del (F267del) variant has been described in two adults with FTD/progressive primary aphasia (32). We initially speculated that all variants in our cohort caused earlier or more profound disruption of cellular function compared to the variants that cause MSP, leading to earlier disease onset. While this could be true for most variants in our cohort, the F267del variant suggests that the syndrome we describe can exist in a spectrum with other *VCP*-related diseases. It would be interesting to know if the adults with this variant had features of the childhood-onset syndrome we describe, or if there are additional risk factors leading to early disease development. The novel variants and phenotypes we describe are distinct from MSP, with no signs of myopathy or bone disease; however, there may be some overlap with other *VCP*-associated neurological conditions. Three probands had abnormal EMG/NCV studies, with evidence of sensorimotor polyneuropathy; these are reminiscent of Charcot-Marie-Tooth type 2Y (MIM #616687), which is caused by VCP variants G97E and E185K (28, 57). Neuropathy appears to be variably penetrant in our cohort, as not all older probands had noticeable neuropathy; formal EMG/NCV studies of more individuals could clarify this association further. Another potential overlapping phenotype is FTD, as the oldest individual in our study, Proband 11, developed atrophy of the frontal lobes on brain MRI. It is possible that specific *VCP* variants primarily impact neurons and spare other cell types, as not all *VCP* variants are associated with neuropathy or dementia. The neurodegenerative effects of *VCP* loss-of-function may be due to accumulation of toxic proteins such as TDP-43 or Tau, as has been shown knockout mice (55) and a VCP hypomorphic variant c.1184A>G (D395G) (58). Additional work is needed to understand if the *VCP* variants in our cohort also produce inclusion bodies, or if there is a novel mechanism leading to early onset of neurological disease. Ongoing evaluation will be important to understand the potential overlap of this new syndrome with other *VCP*-associated syndromes such as MSP, CMT, and FTD.

There are several limitations of this study. Although international collaboration was necessary to compile this cohort of individuals, this meant each proband was evaluated by different healthcare professionals, and analysis was performed with non-standardized sequencing techniques and algorithms. Additionally, there were no uniform measurement tools of the degree of DD/ID, making it more challenging to compare phenotypic severity. Our *in vitro* ATPase studies are necessarily limited, as variants may cause differences in intracellular localization and chaperone binding that are not analyzed in this assay but can drastically affect cellular function. While assessing ATPase function and protein stability provides a point of comparison against MSP-causing variants, more extensive molecular studies are needed to evaluate the impact of the variants in this cohort. For the variants predicted to cause nonsense-mediated decay, functional studies assessing mRNA/protein production, as well as study of behavioral and neurological phenotypes in model organisms, would ideally be performed. We hope the novel variants and phenotype described here will provide rich fuel for future studies and assist in the diagnosis of children with DD/ID.

## CONCLUSION

We present thirteen individuals with a distinct childhood-onset *VCP*-related disorder with developmental delay, intellectual disability, hypotonia, and macrocephaly. We provide evidence that this disease is caused by novel heterozygous variants in *VCP*. Twelve variants were novel and include a frameshift and a splicing variant that could lead to haploinsufficiency, in-frame deletions, and missense variants spread throughout the protein. Additionally, we identify a novel pathomechanism since many of these variants cause decreased ATPase activity and one variant results in hyperactivation of ATPase, whereas the studied MSP-associated variants moderately increase ATPase activity. Overall, this cohort provides a new direction for research and expands our understanding of the functions of *VCP* in neurologic disease.

