## Supplementary materials for "A Novel Autosomal Dominant Childhood-Onset Disorder Associated with Pathogenic Variants in *VCP*"

| ID | mRNA change | Genomic location (chr9) | Protein change | Exon/ domain | Lab class | Allel. freq | Inherit -ance | Effect on prot. | *In silico* | REVEL score | CADD score | Funct. data | ACMG class |
|---|---|---|---|---|---|---|---|---|---|---|---|---|---|
| 1 | c.265del | 35067927 CG>C | p.Arg89Glyfs*8 | 3/N | VUS | PM2 | PS2 | PVS1-M | | | | | LP |
| 2 | c.685C>T | 35064177 G>A | p.Leu229Phe | 6/D1 | VUS | PM2 | PS2* | | PP3 | 0.73 | 4.06/27.5 | PP2 | LP |
| 3 | c.709-2A>G | 35063082 T>C | Splice variant | 6-7/D1 | VUS | PM2 | PS2 | PVS1-M | | | 6.06/35 | | LP |
| 4 | c.753G>T | 35063036 C>A | p.Lys251Asn | 7/D1 | VUS | PM2 | PS2 | | PP3-M | 0.84 | 3.35/24.3 | PP2 | LP |
| 5 | c.766C>G | 35063023 G>C | p.Arg256Gly | 7/D1 | VUS | PM2 | PM6 | | PP3-M | 0.89 | 3.49/24.7 | PP2 | LP |
| 6 | c.801_803del | 35062985 CAAG>C | p.Phe267del | 7/D1 | VUS | PM2 | PS2 | PM4 | | | | | LP |
| 7 | c.812G>A | 35062350 C>T | p.Gly271Asp | 8/D1 | VUS | PM2 | PS2 | | PP3-S | 0.96 | 4.57/32 | PP2 | LP |
| 8 | c.901_903del | 35062258 AGAT>A | p.Ile301del | 8/D1 | VUS | PM2 | PS2 | PM4 | | | | | LP |
| 9 | c.1084C>T | 35061687 G>A | p.Arg362Cys | 10/D1 | VUS | PM2 | PS2 | | PP3-S | 0.98 | 4.80/33 | PP2 | Path |
| 10 | c.1622C>A | 35060386 G>T | p.Ser541Tyr | 14/D2 | VUS | PM2 | PM6 | | PP3-M | 0.86 | 4.18/28.6 | PP2 | LP |
| 11 | c.1874G>C | 35059623 C>G | p.Arg625Pro | 14/D2 | VUS | PM2 | PS2 | | PP3-S | 0.96 | 4.31/29.6 | PP2 | Path |
| 12 | c.2257C>T | 35057434 G>A | p.Arg753Trp | 16/D2 | VUS | PM2 | BS4 | | PP3-M | 0.90 | 4.53/32 | PP2 | VUS |
| 13 | c.892C>T | 35062270 G>A | p.Pro298Ser | 8/D1 | VUS | PM2 | PS2 | | PP3 | 0.71 | 3.94/26.6 | PP2 | LP |

**Supplementary Table 1: *VCP* variant classification**
mRNA change is based on transcript NM_007126.5; genomic location is based on GRCh38.p13 (v1.6, hg38). Classifications of variants in *VCP* use the reporting lab's classification (Lab class) as well as ACMG guidelines (ACMG class) to score pathogenicity based on allele frequency in population databases, inheritance mode/segregation with disease, *in silico* predicted effects on protein (based on REVEL score, but CADD raw/scaled scores also provided), functional data based on mechanism of disease and published data on similar variants in the gene of interest (1-5). CADD score reflects likelihood variant is observed (negative) vs. not observed/only simulated (positive); raw scores are directly from model, scaled scores are compared to all other SNVs and scored by rank order of magnitude (5).

Key for the criteria used:
Benign strong 4 (BS4): non-segregation with disease. Proband 12 had a paternally inherited variant; father with ADHD and possible memory concerns in his 50s, but no report of developmental delay, so considered not affected (and therefore disease not segregating)
Pathogenic supporting 2 (PP2): missense variant in a gene with low ratio of benign to pathogenic missense variants
Pathogenic supporting 3 (PP3): *In silico* predictions with REVEL score of 0.663-0.773; modified to PP3-M (moderate) with REVEL score of 0.773-0.932, and PP3-S (strong) with REVEL score of >0.932
Pathogenic moderate 2 (PM2): none of the variants was reported in gnomAD (last accessed Mar 2023)
Pathogenic moderate 4 (PM4): protein length-changing variant
Pathogenic moderate 5 (PM6): *de novo* without maternity/paternity confirmed (i.e., panel testing)
Pathogenic strong 2 (PS2): *de novo* with maternity/paternity confirmed (i.e., confirmed by trio exome analysis)
Pathogenic very strong (PVS1): predicted null variant in a gene where loss of function is a known mechanism of disease; given haploinsufficiency is not proven mechanism of disease in humans but null mouse model is affected (embryonic lethal)(6), has been downgraded to moderate.
Variant of uncertain significance (VUS): insufficient evidence to label as likely pathogenic or benign
Likely pathogenic (LP): one strong and two moderate/supporting predictions, or ≥3 moderate predictions, among other criteria found in (1)
Pathogenic: two strong predictions, among other criteria found in (1)
* Proband 2/PS2: Half-brother also appears affected, but did not have genetic testing. Not present in parents, so scored as *de novo* here.

| P# | Age (y) | Sex | Pregnancy/birth complications | Relevant family history | Other genetic/metabolic testing |
|---|---|---|---|---|---|
| 1 | 0-5 | M | Low amniotic fluid, brief NICU stay for hypothermia | Maternal half-brother w/suspected autism | Elevated vitamin B12. Normal: carnitine, acylcarnitines, AA, UOA, urine C5-DC, urine GAGs, TSH. Normal CK. |
| 2 | 6-10 | M | Pre-eclampsia, C/S at term | Half-brother with similar features, not tested. | VUS in *TRPC5* (c.2332T>C, p.S778P), maternally inherited. Normal: CMA, UOA, carnitine; Fragile-X testing with 23 repeats. Normal CK, ALP, EMG, muscle ultrasound. |
| 3 | 6-10 | F | Fraternal twin (clomiphene), oligohydramnios, premature C/S, 3mo stay in NICU for poor feeding | Speech delay (both sides); twin with short stature and speech delay | Normal: CMA. ALP elevated to 436 (ref <317) during acute illness. |
| 4 | 11-15 | M | None | 5 healthy siblings, 1 intrauterine fetal demise near term | Normal: CMA, Fragile-X, ANG/PWS MLPA, *ZEB2* seq + MLPA, DDG2P panel. Normal CK, EMG. One normal, one ↑ ALP level. NCV with demyelinating sensorimotor polyneuropathy. |
| 5 | 11-15 | M | None | No additional info | Normal: CMA. |
| 6 | 0-5 | F | C/S at term | No additional info | Normal: karyotype, aCGH. Normal CK and ALP. |
| 7 | 0-5 | F | Mild gDM, C/S at term | Father with small VSD | Normal: karyotype, aCGH, PWS, metabolic screening. Normal CK. |
| 8 | 11-15 | M | Premature C/S for twin pregnancy, choroid plexus bleeding, 2˚ hydronephrosis | Noncontributory | Normal: karyotype, aCGH, *SNRPN* MLPA. Normal CK and ALP. |
| 9 | 16-20 | M | Oligohydramnios, gDM requiring insulin, C/S at term for fetal decelerations | No additional info | Low carnitine with pathogenic variant in *SLC22A5* (p.P46S). Normal: karyotype, Fragile-X, subtelomere FISH, oligo microarray, mtDNA seq. Normal CK, ALP, NCV. EMG with sensorimotor polyneuropathy |
| 10 | 11-15 | F | AMA, term C/S for bradycardia | Noncontributory | Normal: Autism/ID panel, karyotype, Fragile-X, SNP array, ALP. |
| 11 | 21-25 | F | ↓ Fetal movement in 2T, oligohydramnios, vacuum-assisted delivery term, 10d NICU stay for UTI & poor feeding | No additional info | POA suggestive of succinic semialdehyde dehydrogenase deficiency. Normal: karyotype, *CPT-2* seq, Fragile-X, mtDNA panel, CSF, metabolic workup (see P11 supplement). EMG with mixed myopathy/neuropathy, normal 6 years later. Normal ALP. |
| 12 | 16-20 | M | C/S at term | Father with ADHD, memory concerns 50s. | Normal: CMA, metabolic workup (see Proband 10 supplement), mtDNA seq and deletion. Normal ALP. |
| 13 | 11-15 | F | None | Noncontributory | Normal: CMA, sequencing of *SLC2A1* and *STXBP1*, chromosome 15 methylation studies. Normal CK and ALP. |

**Supplementary Table 2: Proband phenotypes (demographics and other contributing factors)**

Key: Age = age at last evaluation/clinical update. Sex: M = male, F = female. Ethnicity: NE = northern European, A = Asian (Thai), H = Hispanic. Pregnancy/birth complications: AMA = advanced maternal age, C/S = Caesarian section, gDM = gestational diabetes mellitus, NICU = neonatal intensive care unit, 2T = second trimester. Other testing: AA = amino acids, aCGH = comparative genomic hybridization array, ALP = alkaline phosphatase, ANG/PWS = Angelman syndrome/Prader-Willi syndrome, CMA = chromosomal microarray, CK = creatinine (phospho)kinase, CSF = cerebrospinal fluid, DDG2P = Developmental Disorders Genotype-to-Phenotype database (>1000 curated genes), EMG = electromyography, GAG = glucosaminoglycan, MLPA = Multiplex Ligation-dependent Probe Amplification, mtDNA = mitochondrial DNA, MPS = mucopolysaccharides, NCV = nerve conduction velocity, POA = plasma organic acids, UOA = urine organic acids, seq = sequencing, SNP = single nucleotide polymorphism, TSH = thyroid stimulating hormone, UTI = urinary tract infection.

| P# | Gross motor delay | | Fine motor delay | | Language delay (first word, age of sentences) | | | Intellectual disability (IQ or clinical estimate) | |
|---|---|---|---|---|---|---|---|---|---|
| Normal | - | Note | - | Reaches at 4mo, pincer grasp 9mo, utensils 2y | - | 1st word | Notes | | IQ > 70 to 75 |
| 1 | +/- | Early milestones mildly late | +/- | Unsteady/slow reach → now age-appropriate | - | - | Age appropriate | - | No concerns |
| 2 | + | | + | Complicated by hypotonia | + | - | | - | Full Scale IQ: 87(low-normal) |
| 3 | + | | + | Difficulty with handwriting, shoelaces, buttons, zippers, and cutting food | + | ++ | 3-word vocab | + | Full Scale IQ: 48 |
| 4 | + | Requires orthoses to walk due to hypotonia, uses wheelchair | + | No additional info | + | ++ | Few words | + | Severe intellectual disability (est 10 years younger) |
| 5 | + | | + | Difficulty writing and cutting food; ↓coordination | + | +/- | Dysarthria | - | School acquisitions slow |
| 6 | + | | - | No additional info | + | + | None | + | Mild-moderate impairment |
| 7 | + | Can lift head, roll, and sit when placed but not independently. Cannot walk. | + | No additional info | + | + | 3-word vocab | + | No additional info |
| 8 | + | Walks with orthosis | + | No additional info | + | - | 5-word vocab | + | Severe impairment |
| 9 | + | Poor balance | + | Poor handwriting | + | - | Dysarthria | + | Mild, 1-3 years below peers |
| 10 | + | | + | Decreased fine motor coordination by OT eval | + | + | Apraxia | + | WISC-V IQ 48 but functionally mild |
| 11 | + | Coordination c/b hemiparesis | + | C/b hemiparesis, tremor, dysdiadochokinesis | + | + | Dysarthria, expressive aphasia | + | IQ in 50s |
| 12 | +/- | | + | C/b tremor of hands | - | Normal speech development | | +/- | IQ 105 → 76 in 6y |
| 13 | + | | + | Tremor and dysmetria | + | + | Limited vocab | + | Developmentally 3y behind |
| **Total** | **13/13** | | **12/13** | | **11/13** | | | **9/13 + 1 drop in IQ** | |

**Supplementary Table 3: Proband phenotypes (developmental delay and intellectual disability)**
Normal milestones based on updated guidelines from CDC and American Association of Pediatrics (7). Any amount of delay was counted. C/b = complicated by.

| P# | Behavioral/psychiatric disorders | | Other neurological disorders | | Abnormal brain MRI | |
|---|---|---|---|---|---|---|
| 1 | - | Brief head-banging behavior, resolved | + | Truncal +/- lower extremity hypotonia, full-body tremor (resolved). Normal EEG. | - | Normal |
| 2 | + | Autism, ADHD, social anxiety, hand-flapping, poor sleep | + | Hypotonia (generalized symmetric limbs), hyporeflexia. Normal EEG. | + | Minimal bilateral frontal, parietal, and periventricular FLAIR changes, 3 years apart |
| 3 | +/- | Some features of autism, hand-flapping behavior | + | Hypotonia | + | Brain MRI as infant w/thin corpus callosum, slightly small pons, slight prominence of extra-axial CSF spaces and ventricles |
| 4 | - | Happy demeanor, hand-flapping behavior | + | Hypotonia (legs), EEG with epileptic activity but no clinical seizures | + | Slightly asymmetric hemispheric sizes, reduced white matter, possible mild polymicrogyria at insulae bilaterally |
| 5 | + | ADHD, anxiety, depression | + | Hypotonia, seizures (on 4 AEDs) | + | Congenital hydrocephalus with quadriventricular dilatation and cerebral atrophy |
| 6 | - | None noted | + | Hypotonia | + | Enlarged perivascular spaces |
| 7 | - | Stereotypic hand movements | + | Hypotonia, two febrile seizures | + | Hydrocephalus, communicating |
| 8 | + | Autism, outbursts, poor sleep | + | Hypotonia | + | Periventricular leukomalacia as infant |
| 9 | + | ADHD | + | Hypotonia, poor coordination/balance | + | Small cranial vault with suggestion of mild global volume loss as adolescent |
| 10 | + | ADHD, anxiety (regarding seizures), outbursts, sleep difficulty | + | Hypotonia (with feeding difficulties in infancy), seizures (on lamotrigine) | + | Mild prominence of lateral/3$^{rd}$ ventricles, mild thinning of corpus callosum/central white matter volume loss |
| 11 | - | None noted | + | Hypotonia, cerebral palsy (R leg hemiparesis/hemiatrophy), coarse action tremor of both hands (on valproate and deep brain stimulator), dysdiadochokinesis | + | Normal initial, mild prominence of extra-axial CSF spaces 6 years later, mild sulcal prominence/ cortical atrophy of frontal lobes 5 years later |
| 12 | + | PDD, autism spectrum, ADHD, anxiety with skin/nail picking, sleep difficulty | + | Bilateral hand tremor, hyperreflexia with clonus, infrequent seizures (AEDs x 4 years, now off) | - | Non-contrast MRI reportedly normal; normal head CT. |
| 13 | - | None noted | + | Hypotonia, action tremor, dysmetria | + | Hypoplasia of corpus callosum splenium, dilation of Virchow-Robin spaces focused in left parietal subcortical area with foliation anomaly and cortico-subcortical dysplasia. |
| **Total** | **6/13 (3 autism spectrum, 5 ADHD, 4 anxiety, 4 sleep)** | | **12/13 hypotonia, 4/13 tremor, 3/13 epilepsy** | | **11/13** | |

**Supplementary Table 4: Proband phenotypes (neurologic and psychiatric disorders)**
Magnetic resonance imaging studies (MRIs) were read at the collaborator's institution. ADHD = attention-deficit/hyperactivity disorder, AED = antiepileptic drug, EEG = electroencephalogram, FLAIR = fluid attention inversion recovery sequence, PDD = pervasive developmental disorder.

| P# | MC | Dysmorphic features | | Congenital anomaly | | Other medical history |
|---|---|---|---|---|---|---|
| 1 | + * | + | Mildly low-set ears, anteverted nares, prominent forehead and chin, positional plagiocephaly | + | Torticollis, inguinal hernia requiring repair, mild hypospadias w/chordee | Pyloric stenosis at s/p pylorotomy, mild thoracolumbar kyphosis |
| 2 | + | + | Broad forehead with frontal bossing, long face, posteriorly rotated ears (L mildly low-set), deep-set eyes, high-arched palate, thin upper lip | + | Bilateral hydrocele | Leg pronation/abnormal hip alignment requiring orthotics, asthma, constipation, joint hypermobility |
| 3 | Rel * | - | Broad forehead, deep-set eyes, bitemporal narrowing, space between upper incisors, thin upper lip | + | Inward-turning feet requiring splinting | GERD, constipation, small kidneys w/hydronephrosis, short stature (2nd percentile) |
| 4 | Rel * | + | DSPF, convex nasal ridge, overhanging nasal tip, uplifted earlobes, thin upper lip, protuberant lower jaw, high palate, long toes | + | Punctate corneal clouding, thin diaphysis/underdeveloped epiphyses of long bones, calcaneovagus deformity, umbilical hernia s/p repair | Scoliosis |
| 5 | + | - | Frontal bossing | + | Congenital hydrocephalus | Astigmatism, hyperopia, GERD |
| 6 | + | + | Dolichocephaly, mild frontal bossing, deep nasal root | - | None noted | Mild myopia |
| 7 | + * | + | Prominent forehead, deep-set eyes, down-slanting PF and eyebrows, low-set/dysplastic ears, long smooth philtrum with thin upper lip, retrognathia, single palmar crease | + | Patent ductus arteriosus, patent foramen ovale/atrial septal defect. | On 4 antihypertensives, strabismus, anisocoria |
| 8 | + | + | Frontal bossing with asymmetry, DSPF, hypotelorism, ptosis, microretrognathia, high palate, dental anomalies | + | Bilateral hip dysplasia requiring surgery, hydronephrosis, cryptorchidism requiring surgery | Strabismus, GERD, gastritis, constipation with encopresis, apneic episodes in infancy |
| 9 | - | - | High foot arches (16-20y), no surgery required | - | None | Scoliosis, mild astigmatism, glasses |
| 10 | + | - | Low-lying columella, thin upper lip | - | None | Thoracolumbar scoliosis, history of feeding difficulties |
| 11 | + | + | Prominent forehead, low-set ears, DSPF, thin upper lip, smooth philtrum; hammer toes, hyperflexible hands/feet, pes cavus | - | None | Ketotic hypoglycemia 0-6y, headaches, constipation, bifocals, skin biopsy w/granular deposits and ↓ eccrine glands,muscle atrophy/cramps, R hemiparesis |
| 12 | + | - | Pes cavus (acquired) s/p surgery | + | Ventricular septal defect | Myopia, constipation |
| 13 | + * | + | Broad forehead, synophrys, DSPF, ptosis, low-set posteriorly rotated ears with simplified folding, full cheeks, smooth philtrum, thin upper lip, microdontia, macroglossia | + | Patent foramen ovale, large left kidney with duplication of urinary system | Many nevi, hyperphagia |
| **Total** | **12/13** | **8/13** | | **9/13** | | |

**Supplementary Table 5: Proband phenotypes (dysmorphic features and congenital defects)**
Facial features evaluated by a geneticist on physical exam; photos also reviewed by corresponding author. Required a significant dysmorphic feature or multiple minor features to be classified as dysmorphic. MC = macrocephaly (+ = absolute, Rel = relative)(8). * Normal OFC at birth with progressive macrocephaly. DS = downslanting, PF = palpebral fissures. GERD = gastroesophageal reflux disease, s/p = status post.

| Proband | 1 | 2 | 3 | 4 | 5 | 6 | 7 | 8 | 9 | 10 | 11 | 12 | 13 | Total |
|---|---|---|---|---|---|---|---|---|---|---|---|---|---|---|
| Mutation | FS | D1 | Splice | D1 | D1 | D1Δ | D1 | D1Δ | D1 | D2 | D2 | D2 | D1 | |
| Sex (M/F) | M | M | F | M | M | F | F | M | M | F | F | M | F | 7/6 |
| Age | 0-5 | 6-10 | 6-10 | 11-15 | 6-10 | 0-5 | 0-5 | 6-10 | 16-20 | 11-15 | 21-25 | 16-20 | 11-15 | |
| ATPase fx | | - | | - | + | + | + | - | - | ++ | - | - | - | |
| | Developmental delay | | | | | | | | | | | | | |
| Gross motor | Mild/- | Mod | Mild | Sev | Mod | Mod | + | Mod | Mild | Mild | Mild | Mild/- | Mild | 13/13 |
| Fine motor | +/- | Mod | + | + | + | - | + | + | + | + | + | + | + | 12/13 |
| Language | - | Mild-Mod | Sev | Sev | + | Mild | Sev | Sev | Mod | Mild | Mild | - | Mod | 11/13 |
| ID | - | + | Sev | Sev | - | Mild-Mod | Sev | Sev | Mild | Mild | Mild-Mod | - (↓IQ) | + | 9/13 |
| | Neurologic & psychiatric disorders | | | | | | | | | | | | | |
| ADHD | - | + | - | - | - | - | - | - | + | + | - | + | - | 4/13 |
| Autism | - | + | +/- | - | - | - | - | + | - | - | - | + | - | 3/13 |
| Anxiety | - | + | - | - | + | - | - | - | - | + | - | + | - | 4/13 |
| Hypotonia | + | + | - | + | + | + | + | + | + | + | + | - | + | 11/13 |
| Epilepsy | - | - | - | - * | + | - | - | - | - | + | - | + | - | 3/13 |
| Tremor | + | - | - | - | - | - | - | - | - | - | + | + | + | 4/13 |
| Abnl MRI | - | + | N/A | + | + | + | + | + | + | + | + | - | + | 11/13 |
| | Dysmorphic features and congenital defects | | | | | | | | | | | | | |
| Dysmorphic | + | + | - | + | - | + | + | + | - | - | + | - | + | 8/13 |
| Macrocephaly | + | + | Rel | Rel | + | + | + | + | - | + | + | + | + | 11/13 |
| MSK | + | + | + | + | - | - | - | + | + | + | + | + | - | 9/13 |
| Ophthal. | - | - | - | + | + | + | + | + | + | - | + | + | - | 8/13 |
| GI | + | + | + | - | + | - | - | + | - | + | + | + | - | 8/13 |
| GU | + | + | - | - | - | - | - | + | - | - | - | - | + | 4/13 |
| Cardiac | - | - | - | - | - | - | + | - | - | - | - | + | + | 3/13 |

**Supplementary Table 6: Summarized proband phenotypes**

Aggregated phenotypic data for the thirteen probands in our cohort. For mutations: FS = frame shift, D1Δ = in-frame deletion in D1, D1 or D2 = missense in this domain. ATPase function was increased (+) or decreased (-) compared to wildtype (see Figure 2C). Macrocephaly was absolute (+) or relative (Rel). For the purposes of this table, we have qualified developmental delay as: mild delay of ≤2x months to milestone, mod (moderate) >2x normal, sev (severe) = never reached milestone. For ID, based on IQ score < 75 and/or clinical impression, with mild IQ 55-75, moderate 35-55, and severe <35. Musculoskeletal (MSK) abnormalities included kyphosis, scoliosis, hammer toes, pes cavus, long bone deformities, and hip or foot dysplasias, deformities, or misalignments. Ophthalmologic abnormalities include corneal clouding, astigmatism, strabismus, and hyperopia/myopia. Gastrointestinal (GI) abnormalities include pyloric stenosis, reflux, constipation, and poor feeding. Genitourinary (GU) abnormalities include hypospadias, cryptorchidism, hydrocele, and duplicated collecting system. Cardiac abnormalities include atrial and ventricular septal defects and patent ductus arteriosus. *Proband 4: EEG with epileptic activity but no clinical seizures.

**SUPPLEMENTARY MATERIALS**

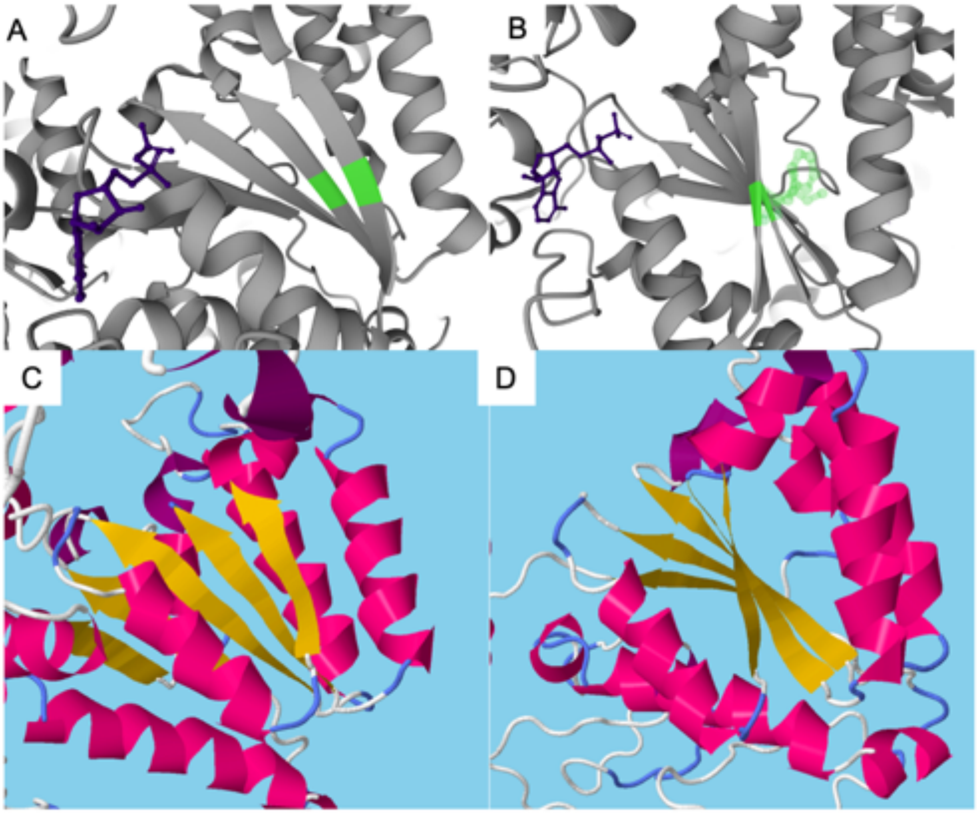


**Supplementary Figure 1: 3D structure of variants of Probands 6 (F267del) and 8 (I130del).** A, B) Two views of a section of VCP, with the F267 and I301 residues highlighted in green, and ADP in purple. 3D models are based on electron microscopy of VCP ADP-bound hexamer (7BP9, viewed in Uniprot browser)(64). In B), the phenylalanine and isoleucine residues are shown in ghost form, interacting with a nearby alpha helix that helps form the “top” surface of the hexamer. The same section of VCP is shown from modeling of the C-D) F267del and E-F) I301del variants, as predicted by the software RaptorX (37, 38). The overall secondary structure is roughly maintained, with no striking displacements of the nearby alpha helices.

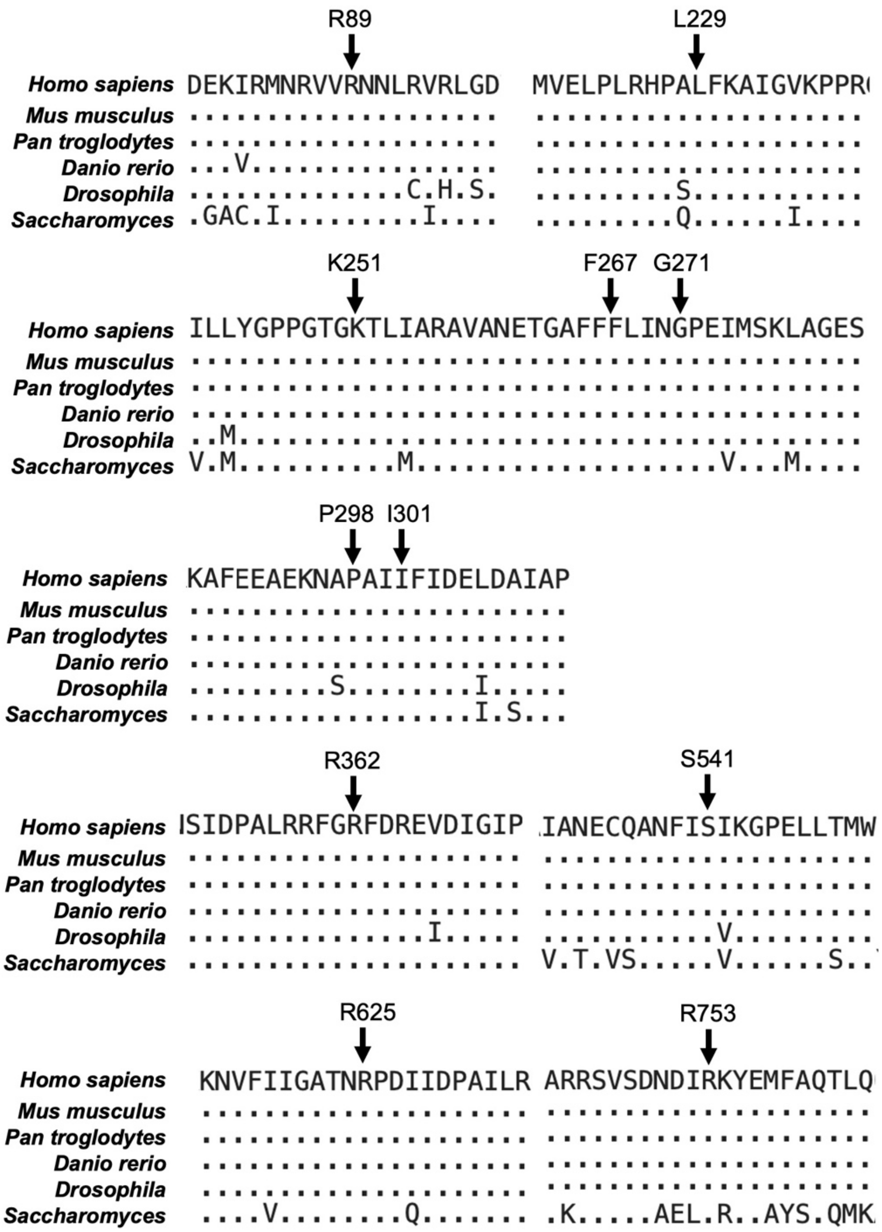


**Supplementary Figure 2: Conservation of affected VCP residues**.
NCBI BLAST was used to align VCP protein sequences from human (*Homo sapiens*) against five model organisms: chimpanzee (*Pan troglodytes*), zebrafish (*Danio rerio*), fruit fly (*Drosophila melanogaster*), and yeast (*Saccharomyces cerevisiae*). Conserved residues are shown by dots, and variant amino acids are shown at each position. Positions affected in our probands are highlighted along with 10 amino acids on either side. Notably, every residue affected is conserved in these organisms, and most are in highly conserved areas.

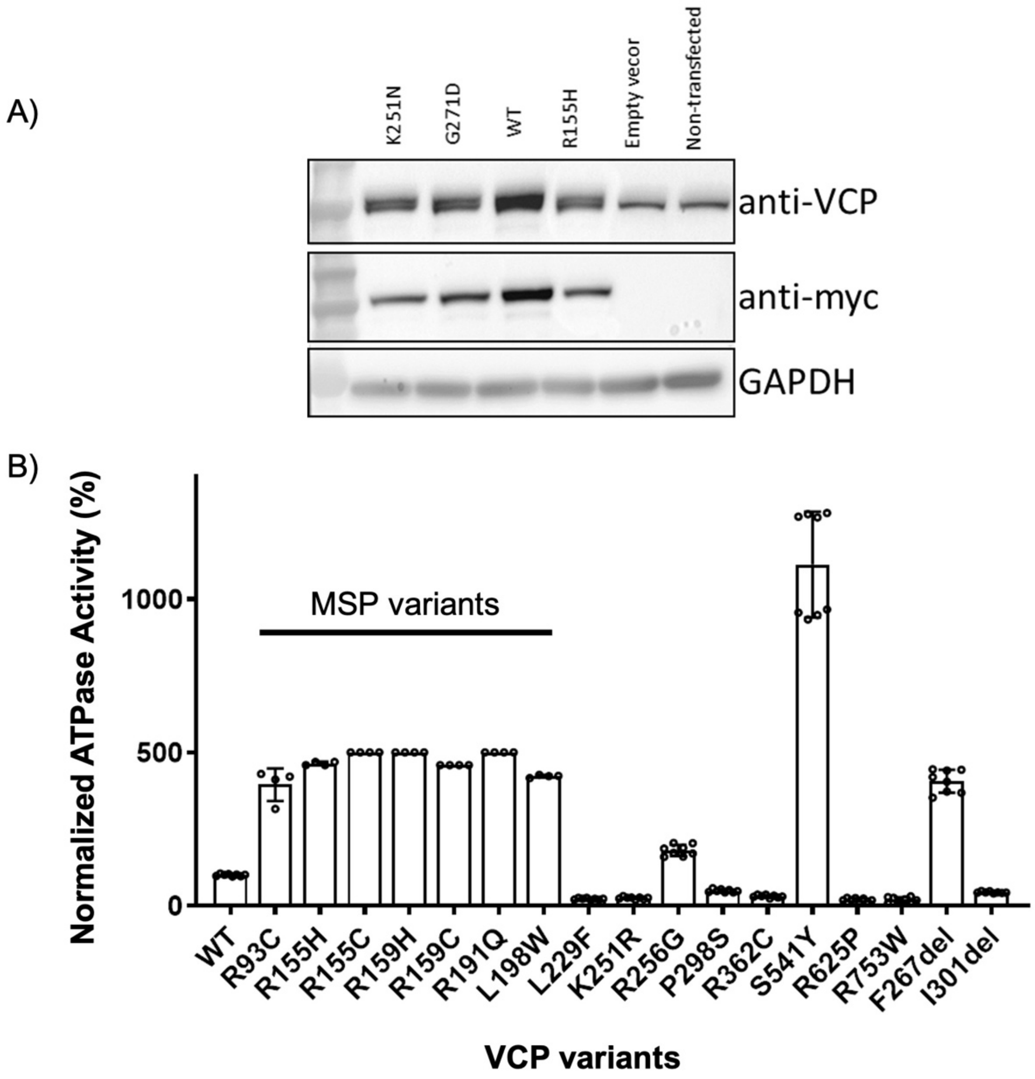


**Supplementary Figure 3: ATPase activity compared to wildtype and MSP variants**
A) Immunoblots of lysates from U2OS cells expressing empty vector or expression plasmids with *VCP* variants and a C-terminal Myc tag using an anti-VCP antibody (top), anti-Myc antibody (middle) or anti-GAPDH as loading control (bottom). B) *In vitro* constructs using myc-tagged VCP were created using variants described in classic multisystem proteinopathy ("MSP variants") and with the novel variants described in this paper. These constructs were transfected into U2OS cells that contained endogenous VCP, and ATPase activity was measured from cell lysates using standard colorimetric assays as in Figure 2C. Activity was normalized to wildtype (100%). In general, MSP variants have up to 5x increased ATPase activity, while most novel variants (with the exceptions of R256G, S541Y, and F267del) had decreased activity.

**PROBAND INFORMATION**
Please contact corresponding author if you are interested in more detailed clinical information about our probands.

**Proband 1**
VCP variant: c.265del (p.R89Gfs*8)

Genetic Testing: Trio whole exome sequencing identified a *de novo* frameshift alteration in *VCP* (c.265del; p.Arg89GlyfsTer8) and no other reportable variants were identified.

Methods: Standard laboratory procedures were followed for whole exome trio analysis performed in a diagnostic laboratory. Briefly, Agilent SureSelectXT Clinical Research Exome kit was used to target known disease-associated exonic regions of the genome and sequenced with Illumina NovaSeq$^{TM}$ 6000 System with 150 bp paired-end reads. Data were aligned with Illumina DRAGEN Bio-IT Platform software (hg19/NCBI build 37) and the emedgene software (Illumina) was used to filter and analyze variants.

**Proband 2**
VCP variant: c.685C>T (p.L229F)

Genetic Testing: Trio whole exome sequencing revealed two VUS; a *de novo* mutation in *VCP* (c.685C>T, p.L229F), and a maternally inherited mutation in *TRPC5* (c.2332T>C, p.S778P). Other testing included normal CMA in 2014 and a 23 CGG repeat on Fragile X testing. The proband was enrolled in the SPARK study for autism; no additional genetic variants were found.

Methods: DNA was isolated from the proband and his parent's blood samples or cheek swabs using standard methods and sent to GeneDx for trio whole exome sequencing. Analysis from GeneDx used their usual protocol: an Illumina system was used to obtain sequences which were then aligned to human genome build GRCh37/UCSC hg19 and analyzed using Xome Analyzer. Pathogenic variants were confirmed by capillary sequencing or other appropriate method, and variants in the proband were compared to parental samples. Re-analysis by GeneDx in 2019 did not change their classification or report any additional variants.

**Proband 3**
VCP variant: c.709-2A>G (splice variant)

Genetic Testing: Trio whole exome sequencing was performed, which found a *VCP* c.709-2A>G splice variant. She also had a normal chromosomal microarray.

Methods: DNA was isolated from the proband and her parent's blood samples or cheek swabs using standard methods and sent to GeneDx for trio whole exome sequencing. Analysis from GeneDx used their usual protocol, as for Proband 2. The variant found was reported as a de novo variant of uncertain significance.

**Proband 4**
VCP variant: c.753G>T (p.K251R)

Genetic Testing: A *de novo* VCP variant, c.753G>T (p.Lys251Arg), was identified by trio whole exome sequencing. Testing with normal results performed prior to exome sequencing included: Chromosomal microarray, *FMR1* sequencing, MLPA for Angelman/Prader Willi syndrome,

MLPA and sequencing for *ZEB2*, and a developmental disorders gene sequencing panel (DDG2P, contains 1641 genes, performed in 2017). No other genetic lesion was found.

Methods: Trio whole-exome analysis was performed at Telemark Hospital Trust. Nextera Rapid Capture Exome Enrichment Kit (Illumina) was used for sample preparation and a NextSeq 500 (Illumina) for 2x150 bp sequencing. Reads were mapped to the reference genome (GRCh37/hg19) by BWA (Burrows-Wheeler Aligner)(12). GATK (Genome Analysis Toolkit) was used for variant calling (13). Variants were annotated by Annovar (14) and filtered with Filtus (15).

**Proband 5**
VCP variant: c.766C>G (p.R256G)

Genetic Testing: Gene panel testing detected a *de novo* c.766C>G, p.R256G variant in *VCP*. Chromosomal microarray was normal.

Methods: Sequencing was performed at the Molecular Genetics Laboratory at Brest University Hospital. The panel was comprised of ~500 genes associated with intellectual disabilities. A parental pool strategy was used, and variant above was confirmed to have *de novo* segregation by Sanger sequencing.

**Proband 6**
VCP variant: c.801_803del (p.F267del)

Genetic Testing: Trio exome sequence detected a de novo variant in VCP, c.801_803del. She had chromosome analysis and array CGH with normal results. This variant was recently reported (Feb 2023) as a VUS in ClinVar (variation ID 1913089) (Last accessed March 2023).

Methods: Trio whole exome sequencing (WES) was performed at the Institute of Human Genetics, Helmholtz Center Munich. Exome-sequencing was performed with a SOLiD 5500XL machine (Life Technologies) after enrichment with the Agilent SureSelectXT Human All Exon 50Mb Kit (Agilent). The data were analyzed with LifeScope software (Applied Biosystems, Life Technologies). Trio-WES data were filtered for potentially pathogenic *de novo* variants absent in the general population (dbSNP138, 1000 Genomes Project, and gnomAD Browser) and rare biallelic variants with a minor allele frequency (MAF) <0.1% and no homozygous carriers in the aforementioned databases. The functional impact of identified putative pathogenic variants was predicted by the Combined Annotation Dependent Depletion (CADD) tool (5), the Rare Exome Variant Ensemble Learner (REVEL) scoring system (4), the Mendelian Clinically Applicable Pathogenicity (M-CAP) Score (16), the Sorting Intolerant from Tolerant (SIFT) program (17, 18), and the Polymorphism Phenotyping v.2 (PolyPhen-2) tool (19).

**Proband 7**
VCP variant: c.812G>A (p.G271D)

Genetic Testing: The *VCP* c.812G>A variant was found on trio exome sequencing. This was classified as a *de novo* variant of uncertain significance by the sequencing lab. She previously had a chromosome analysis, array CGH, and Prader-Willi syndrome methylation analysis that were normal. Reportedly, metabolic screening tests on her 2$^{nd}$ day of life that were also unremarkable.

Methods: Written informed consent for sequencing and research was obtained in accordance with the German gene diagnostics act and the declaration of Helsinki. Sequencing was performed at the laboratory of Charité Universitätsmedizin Berlin. Genomic DNA was extracted from whole blood samples and exome enrichment performed using the SureSelect Human All Exon Kit V6 (Agilent Technologies). Samples were sequenced on an Illumina NovaSeq6000 in 2x100bp paired-end mode. Reads were mapped to the human reference genome GRCh37/hg19 and variants filtered by minor allele frequency, mode of inheritance and predicted impact using the VarFish platform (20).

**Proband 8**

VCP variant: c.901_903del (p.I301del)

Genetic Testing: Trio whole exome sequencing was performed and a *de novo* variant in *VCP* [NM_007126.5:c.901_903del, p.(Ile301del)] was identified in the proband. No other notable variants were reported. Prior negative genetic workup included MLPA for the SNRPN locus, karyotyping, and array CGH.

Methods: Trio whole exome sequencing was performed at the D&R Institute for Human Genetics, Medical University of Graz. DNA was extracted using the QIASymphony DSP DNA Midi Kit on a QIASymphony SP instrument (QIAGEN, Hilden, Germany). Nextera DNA Flex Library Prep Kit was used for library preparation. Sequencing was performed on a NextSeq 550 (Illumina, San Diego, California, USA). Alignment to the human reference sequence (GRCh37/hg19 assembly) and variant calling were performed with the DRAGEN Germline Pipeline V.3.2.8 on Illumina BaseSpace (https://basespace.illumina.com/). Variant annotation and filtering was done via VarSeq™ v 2.2.2 (Golden Helix, Inc., Montana, UCS, www.goldenhelix.com). For variant prioritization following human phenotype ontology (HPO) terms were used: HP:0001263 Global developmental delay, HP:0100716 Self-injurious behavior, HP:0000729 Autistic behavior, HP:0006970 Periventricular leukomalacia, HP:0025069 Concomitant strabismus, HP:0020045 Esodeviation, HP:0001252 Muscular hypotonia, HP:0002705 High, narrow palate, HP:0001385 Hip dysplasia, HP:0000028 Cryptorchidism, HP:0000256 Macrocephaly, HP:0002007 Frontal bossing, HP:0000494 Downslanted palpebral fissures, HP:0000601 Hypotelorism, HP:0005656 Positional foot deformity, HP:0000308 Microretrognathia, HP:0005949 Apneic episodes in infancy.

**Proband 9**

VCP variant: c.1084C>T (p.R362C)

Genetic Testing: Trio exome sequencing revealed a single reported finding, a c.10842C>T (p.R362C) *de novo* variant of uncertain significance in *VCP*. Mitochondrial DNA sequencing did not find any pathogenic variants. Unremarkable karyotype, subtelomeric FISH, oligo microarray, and Fragile-X repeat analysis. *SLC22A5* pathogenic variant discussed above.

Methods: Genomic DNA from the submitted proband was enriched for the complete coding regions and splice site junctions for most genes of the human genome using a proprietary capture system developed by GeneDx for next-generation sequencing with CNV calling (NGS-CNV). Data were aligned to reference sequences based on NCBI RefSeq transcripts and human genome build GRCh37/UCSC hg19 and analyzed to identify sequence variants and most deletions and duplications involving three or more coding exons.

**Proband 10**
VCP variant: c.1622C>A (p.S541Y)

Genetic Testing: GeneDx's Autism/ID Xpanded panel identified a c.1622C>A (p.S541Y) variant in *VCP*. No other variants were reported. Prior testing includes karyotype, SNP microarray, and Fragile-X analysis which were normal.

Methods: Genomic DNA from the proband and both parents was analyzed using next-generation sequencing with CNV calling. Analysis was targeted a phenotypic-driven gene list (GeneDX Autism/ID Xpanded panel). Data were aligned to reference sequences based on NCBI RefSeq transcripts and human genome build 10 GRCh37/UCSC hg19 and analyzed to identify sequence variants and most deletions and duplications involving three or more coding exons.

**Proband 11**
VCP variant: c.1874G>C (p.R625P)

Genetic Testing: Whole exome sequencing showed a heterozygous *de novo* variant in *VCP* (c.1874 G>C), classified as "variant, likely mutation." The following were normal: *ATM*, *CDKL5/STK9, DYT1*, *MeCP2, PEO1*, *POLG1,* partial CMT-axonal DNA testing, Friedreich ataxia, Fragile X, mtDNA point mutation and deletion panel, and chromosomal 15 PCR test.

Methods: Trio exome sequencing was performed at GeneDx on blood from the proband and parents using Agilent SureSelect XT2 All Exon V4 kit, the Illumina 2000 sequencing system (214X mean depth coverage), mapped to the human genome build UCSC hg19 reference.

**Proband 12**
VCP variant: c.2257C>T

Genetic Testing: Trio whole exome sequencing identified a paternally inherited variant of uncertain significance in *VCP*, c.2257C>T (p.R753W). It was reanalyzed as an adult which did not report any new variants or secondary findings. He also had an unremarkable chromosomal microarray.

Methods: Trio whole exome sequencing and re-analysis was performed at GeneDx, using their usual protocols.

**Proband 13**
VCP variant: c.892C>T (p.P298S)

Genetic Testing: Previous workup was normal and included CMA, sequencing of *SLC2A1* and *STXBP1*, and chromosome 15 methylation studies.

Methods: Not available for this proband.
